# Mapping the plague through natural language processing

**DOI:** 10.1101/2021.04.27.21256212

**Authors:** Fabienne Krauer, Boris V. Schmid

## Abstract

Pandemic diseases such as plague have produced a vast amount of literature providing information about the spatiotemporal extent of past epidemics, circumstances of transmission, symptoms, or countermeasures. However, the manual extraction of such information from running text is a tedious process, and much of this information has therefore remained locked into a narrative format. Natural Language processing (NLP) is a promising tool for the automated extraction of epidemiological data from texts, and can facilitate the establishment of datasets. In this paper, we explore the utility of NLP to assist in the creation of a plague outbreak dataset. We first produced a gold standard list of toponyms by manual annotation of a German plague treatise published by Sticker in 1908. We then investigated the performance of five pre-trained NLP libraries (Google NLP, Stanford CoreNLP, spaCy, germaNER and Geoparser.io) for the automated extraction of location data from a compared to the gold standard. Of all tested algorithms, spaCy performed best (sensitivity 0.92, F1 score 0.83), followed closely by Stanford CoreNLP (sensitivity 0.81, F1 score 0.87). Google NLP had a slightly lower performance (F1 score 0.72, sensitivity 0.78). Geoparser and germaNER had a poor sensitivity (0.41 and 0.61) From the gold standard list we produced a plague dataset by linking dates and outbreak places with GIS coordinates. We then evaluated how well automated geocoding services such as Google geocoding, Geonames and Geoparser located these outbreaks correctly. All geocoding services performed poorly and returned the correct GIS information only in 60.4%, 52.7% and 33.8% of all cases. The rate of correct matches was particularly low when it came to historical regions and places. Finally, we compared our newly digitized plague dataset to a re-digitized version of the plague treatise by Biraben and provide an update of the spatio-temporal extent of the second pandemic plague outbreaks. We conclude that NLP tools have their limitations, but they are potentially useful to accelerate the collection of data and the generation of a global plague outbreak database.

## Introduction

Information about the places and times of epidemics are among the core aspects of infectious disease epidemiology. One of the most notorious infectious diseases – the plague – has produced a large body of publications about its historical spatio-temporal spread. Among the most complete compilations of places where plague reportedly occurred are the works of Sticker in 1908 (Sticker, 1908) and Biraben in 1975 (Biraben, 1975). A brief overview over other publications is given in Table S1. Narrative plague texts must typically be converted into quantitative data in order to be usable for epidemiological analyses. However, the manual extraction of data from running text is time and labor intensive.

In the past few years, advances in machine learning algorithms and increasing computing efficiency have led to a rise of digital methods in epidemiology (Salathe, 2018). Particularly the automated generation of data from text through Natural Language Processing (NLP) has gained popularity. For example, NLP approaches have been used to analyze the spread of infectious diseases based on social media postings (Broniatowski et al., 2013) or to analyze the geographical distribution of cholera mentions in the UK Registrar General’s reports from England and Wales in the 19^th^ century (Murrieta-Flores et al., 2015). The plague dot txt project at the University of Edinburgh has recently started to develop a NLP workflow to build a structured account of plague epidemiology based on treatises and publications about the third pandemic (Casey et al., 2020). To our knowledge, the latter is the only project to date that explores the use of NLP in plague research.

The possibilities of NLP algorithms are manifold. They can partition a text word-wise (tokenization), analyze the syntax (position-of-speech, POS), identify entities (named entity recognition, NER) or analyze the sentiment or relations among entities. The NER analysis identifies and classifies tokens into pre-defined categories based on rules (i.e. a dictionary), statistical predictions, or both. A special case of NLP NER is the extraction of geographical data from a text (geoparsing). Geoparsing comprises the identification of a geographical entity (toponym) and the linkage of the geographical entity with GIS data such as coordinates (geocoding). In theory, both steps can be done by hand and/or separately, but automated workflows may be preferable because they are faster and more reproducible.

In general, text mining tools can accelerate the generation of large spatio-temporal datasets, but their performance has to be sufficient to outweigh the errors arising from the automated process. The performance of these algorithms depends on the chosen model or algorithm, and the structure and language of the text. Ideally, an NLP algorithm has a high recall or sensitivity (e. g. the proportion of locations that are correctly identified as locations) and a high specificity (e. g. the proportion of non-locations that are correctly identified as non-locations). Various NLP algorithms and libraries have been tested for modern English medical and non-medical texts and their performances differ substantially (see e.g. (Dreisbach et al., 2019; Gritta et al., 2018)). The sensitivity and the precision of the Edinburgh Geoparser, a popular tool for historical English texts, was found to vary between 60 and 80% depending on the text type and the relative frequencies of the location entities (Grover et al., 2010).

There is a growing scientific interest in building a global database of historical plague outbreak (van Bavel et al., 2019). The Black Death Digital Archives project (http://globalmiddleages.org/project/black-death-digital-archive-project) initiated by Green and Roosen aims to “newly interrogate our traditional sources of historical information” and to link biological, archaeological and documentary databases (Green and Roosen, 2019). We here contribute to this effort with a case study on the use of NLP to facilitate the digitization of plague location data. We use the plague treatise by Sticker (Sticker, 1908) as an example. We compare the application of different NLP libraries and geocoding services for the extraction of place names and coordinates. Finally, we compared our novel, geocoded plague dataset to Biraben’s plague dataset - which we newly re-digitized and geocoded - to highlight the benefits of drawing information from a broader corpus of literature.

## Methods

### Source text

A short description of the structure of Sticker’s work is given in the supplement (Text S1). The text is a combination of running text interspersed with semi-tabulated year and place listings. The running text contains both specific information about places that mention plague in a given year, but also general information on plague as well as historical anecdotes and elaborations. A scanned OCR version of the book is freely available on the Internet Archive (https://archive.org/details/abhandlungenausd01stic/mode/2up).

### Pre-processing and establishment of gold standard location list

In a first preprocessing step, we cleaned the raw OCR text manually. We removed interspersed tables, end-of-line hyphenations, page numbers, headers, and notes in the book margins. We corrected misaligned text and checked the text file for OCR errors by looking for special characters and words that were not recognized by the Notepad++ Spell Checker. To facilitate the automated geoparsing approach, we also removed all words or sentences in parentheses, which were mainly author names and references and thus irrelevant for the tagging. We then established the gold standard list of location toponyms, with both authors independently annotating the preprocessed text using the annotator tool webanno (version 3.5.9) (Eckart de Castilho et al., 2016). We included all administrative place, region or country names as well as natural features such as “the Black Sea”. Associative toponyms such as “the Bishop of Avignon” were excluded because they are not true locations. We then compared our two annotations and established a consensus document. This list of toponyms contained all geographical entities in the text irrespective of whether the location was linked to plague or not. This gold standard list was used for the evaluation of the tagging performance of various NLP libraries (see below). A schematic of the workflow is shown in Fig. S1.

### Establishment of plague dataset

We used the gold standard location list to generate the final dataset of places with plague outbreaks. For this we extracted text snippets of 100 characters before and after each toponym to obtain the context and decided for each case individually whether it was linked to a specific plague outbreak. If the context was unclear, we referred back to the original text. Furthermore, we extracted the corresponding years (usually a four-digit string) using regular expression (regex) and allocated them manually to the corresponding toponym. We also linked the referenced author names (i.e. the source of the information) with the corresponding places wherever it was available. Finally, we batch geocoded the locations of the plague dataset using the REST API services of ArcGIS (https://developers.arcgis.com/rest/) to query the GIS information for each place. We extracted the modern place names, the country ISO code, the centroid and bounding box coordinates and the type of administrative unit. The bounding box coordinates are the minimum and maximum longitudes and latitudes of a given administrative unit, and can be used a proxy for the spatial extent of a place. The coordinates are provided in WGS84. All ArcGIS geocoded locations were individually inspected and mapped to detect improbable results. Ambiguous or unclear toponyms or questionable results were checked individually by consulting the original literature or other sources referenced therein. Entries that could not be identified through automatic geocoding were looked up and coded manually if identifiable. Historical or colloquial regions without a clear administrative border were geocoded approximatively by defining the boundary coordinates manually based on maps on Wikipedia and calculating the arithmetic centroid coordinates. Toponyms that could not be localized exactly were geocoded according to the next lower identifiable level administrative unit and were marked as approximate. Toponyms that could not be localized at all were marked as unknown. We categorized all results as one of the following: place (city, town, village, neighborhood, district, municipality and other populated place), administrative unit (county, state and province), country, island and region (colloquial area, historical or geographical region, and natural features such as streams, mountains or lakes). This dataset was used for the performance evaluation of the geocoding algorithms and is also the final output of our study. The study was conducted in a Windows environment with a german locale. All work was carried out in R/R Studio (version 4.0.0) and Notepad++. The R code and the final plague datasets are available in a repository (https://doi.org/10.5281/zenodo.6587267) (Krauer and Schmid, 2021).

### Toponym NER performance evaluation

We tested four different NLP libraries and one geoparser for the identification of toponyms: Google NLP (Google Ireland Limited, 2019a), Stanford CoreNLP (Manning et al., 2014) with the pre-trained German model version 2018-10-05 (Faruqui and Padó, 2010), spaCy (Explosion, 2019b) with the pre-trained German model version 2.1.0 (Explosion, 2019a), germaNER (Benikova et al., 2015) and Geoparser.io (Geoparser Inc, 2019). For a technical comparison of the libraries see supplement Table S2. We performed syntax analysis (POS) to obtain the tokens, and named entity recognition (NER) to obtain the toponyms. All libraries except germaNER require running text as input. The NER for germaNER was done using the tokenization returned by spaCy. Geoparser.io only returns toponyms and the corresponding GIS information but not the tokenization of the complete text. Google NLP, spaCy and Stanford CoreNLP each have a different algorithm for tokenization, which results in a slightly different numbers of tokens returned. The main difference arises from how the different libraries treat punctuation in relation to words or numbers (e.g. “usw.” or “1346-47” may be treated as one or two tokens). Google, Stanford coreNLP and geoparser do not accept pre-tokenized text as input. For an accurate comparison we combined all in one dataset by mapping all the entities to the tokens of the gold standard. After the mapping, we re-categorized the entities of all five approaches as “location” or “other” (which includes non-identified tokens). If geographical entities were not recognized completely by a text mining algorithm, we allowed also for partial matches for the calculation of the performance indicators. For example, “Freiburg im Breisgau” could be identified as “Freiburg” or the full name. We then compared the sensitivity (recall, true positive rate), the specificity (selectivity, true negative rate), the accuracy, the positive and negative predictive value (PPV and NPV), the F1 score and Cohen’s Kappa. The formal definition of all measures is given in supplement Text S2 and Table S3. The German Stanford CoreNLP java library (version 2018-10-05) was downloaded from the Stanford NLP Github Page (https://stanfordnlp.github.io/CoreNLP/human-languages.html) and accessed through the R package coreNLP (version 0.4.2) (Arnold and Tilton, 2016). SpaCy (v2.0) was downloaded and accessed through the R package spacyr (version 1.2) (Benoit and Matsuo, 2019). The java standalone for GermaNER was downloaded from the Github account (https://github.com/tudarmstadt-lt/GermaNER) and run from the command line.

### Geocoding performance evaluation

We also assessed the performance of three alternative geocoding services: Google (Google Ireland Limited, 2019b), Geoparser.io (Geoparser Inc, 2019), which combines the toponym recognition and geocoding, and Geonames (GeoNames, 2019). Geoparser.io returns only the name of the toponym, the type and the centroid coordinates. Google and Geonames.io provide more GIS information such as lower level administrative area units and place names in local or alternative languages. Google and Geoparser.io return the best match (according to internal criteria), while Geonames returns all possible matches in a ranked order. To make the algorithms comparable we picked only the first (i.e. best) match returned by Geonames. However, we restricted the Geonames search to places (P), administrative units (A), areas (L) and natural features (T, H and V). If no full match was found, we accepted also partial (fuzzy) match for Geonames and Google. We defined the following conditions for a result to be a match: 1) If both the gold standard entity and the comparator entity were a country and the country ISO codes agreed, 2) If both the gold standard entity and the comparator entity were a place or region in the same country and the Euclidian distance between the centroids of the standard and comparator was less than 30 km (for small entities with a standard bounding box up to 30 km), or less than half of the bounding box diameter of the standard (for larger entities with a standard bounding box diameter of more than 30 km). Based on the count of matches we calculated the proportion of toponyms identified (i.e. whether there was a result nor not) and the proportion of toponyms correctly identified for each approach. We also examined the mismatches and checked whether there was a potential regional or other bias in the geocoding. All Geocoding services were accessed through their REST-APIs between September and October 2019 using a designated batch geocoding script.

### Plague data description and comparison

Finally, we summarized the spatial and temporal coverage of our dataset and compared it with a re-digitized version of Biraben’s list (see supplemental Text S2). For this, we merge the two datasets by year and centroid coordinates. We calculated the proportion of full matches among all observations of both datasets for the same time period, plotted all locations in both datasets and compared the corresponding time series. We then restricted the merged dataset to the time period of the second pandemic and to exactly localized places (without regions, countries or other administrative areas) to update and summarize the spatio-temporal extent of the plague outbreaks.

## Results

### Gold standard

The cleaned OCR text of chapters five to sixteen of Sticker’s treatise on plague was 864,106 characters long. Removing the author citations that are present throughout the text reduced the length to 842,918 characters. We identified 7884 geographical entities (5.4% of all tokens) with manual annotation. Of these 7884 toponyms only 4474 (57%) referred to a specific plague outbreak in a specific year (Fig. 1). The rest were mainly repeated mentions of the same locations for a given year or additional geographical information to describe a place (e.g. “Geverske *near* Ostrovizza *in the region of* Zara”). Of these 4474 toponyms, 4087 (91.4%) could be localized exactly. Eight toponyms (0.2%) could not be localized at all (“unknown”). The remaining 379 toponyms (8.5%) were either colloquial or historical regions (e.g. “Podolia”) without clearly defined modern boundaries, or populated places that could not be localized exactly but were attributed to a lower level administrative unit. These are marked as “approximate” in the final dataset. The automated geocoding procedure matched 93.8% of all entries, but 6.2% could not be geocoded with ArcGIS and had to be looked up manually. Only 4.8% of all entries that were historical regions and 21% of all entries that were colloquial areas could be identified automatically through the ArcGIS geocoding services.

**Fig. 1.**
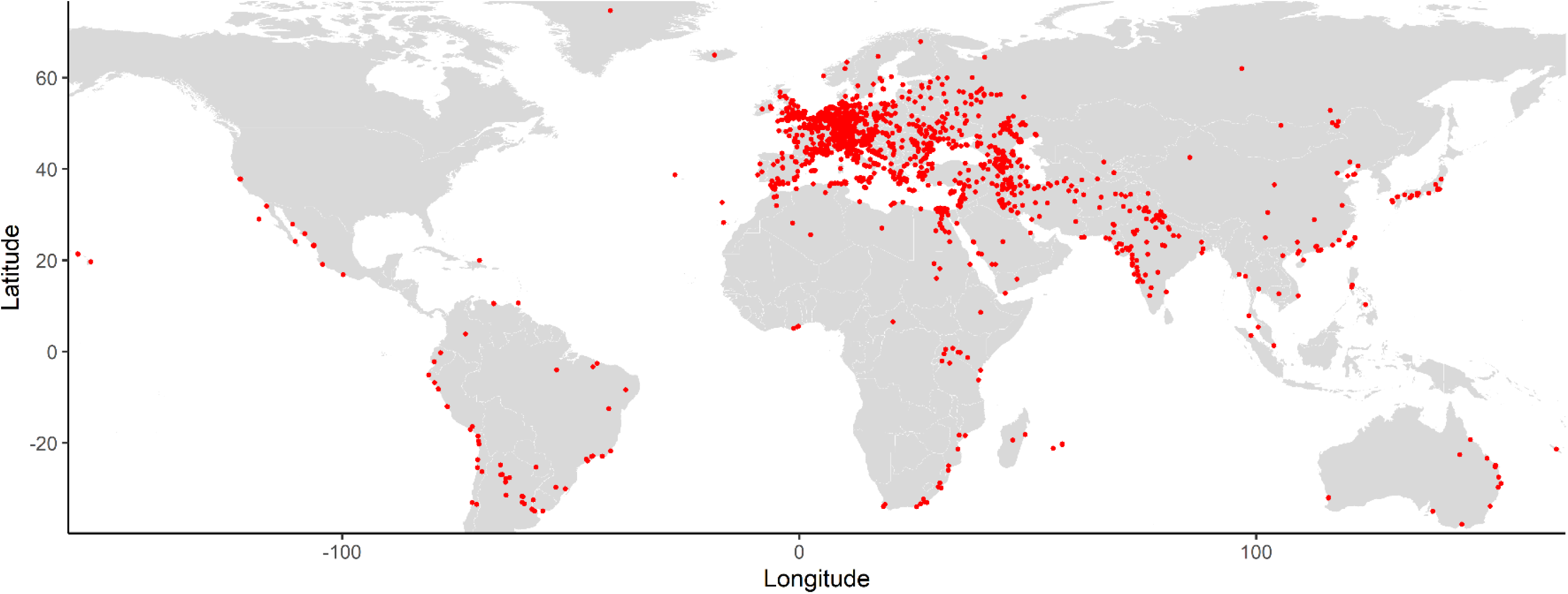
Spatial coverage of all geocoded geographical units (exact and approximate). Note that the data include different administrative levels from village to country, and the dots denote the centroids of each geographical entity.

### Toponym NER performance evaluation

The spaCy and the Stanford CoreNLP tokenizers yielded a similar number of total tokens (146,766 and 146,743 respectively) while Google NLP returned marginally less tokens (146,340) (Table S4). GermaNER identified the most entities (34% of all recognized tokens = 50,374), followed by Google NLP (23% of all recognized tokens = 33,925), spaCy (9% of all recognized tokens = 12,963), Stanford coreNLP (6.5% of all recognized tokens = 9522) and Geoparser.io (3563). After mapping the results to the standard tokenization, Google and spaCy identified the largest percentage of all tokens identified as locations (6.4% and 6.6%), followed by Stanford coreNLP (4.6%) and germaNER (3.9%). The Geoparser.io algorithm identified only 2.4% of the tokens as locations.

Overall, the proportion of correctly identified entities (accuracy) was large for all five libraries (range 0.97-0.99) (Table 1). The spaCy library showed the highest sensitivity (0.92), followed by Stanford CoreNLP (0.82), Google (0.78), germanNER (0.62) and Geoparser.io (0.41). The specificity was equally high for all algorithms (range 0.98-0.99). Stanford coreNLP had the highest precision (PPV, 0.95, i.e. 5% of the positives are false), followed by Geoparser.io (0.9), germaNER (0.85), spaCy (0.75) and Google (0.66). The F1 scores and Cohen’s kappa coefficients suggested a good overall performance for Stanford CoreNLP (0.88 and 0.87) and spaCy (0.83 and 0.81), a mediocre overall performance for Google NLP (0.72 and 0.70) and GermanER (0.71 and 0.70) and a poor performance for Geoparser.io (0.56 and 0.55).

**Table 1.** Performance of different NLP algorithms for the identification of toponyms (location nouns) after mapping to a common tokenization.

|  | Google NLP | Stanford<br>CoreNLP | spaCy | germaNER | Geoparser |
| --- | --- | --- | --- | --- | --- |
| TP | 6154 | 6435 | 7267 | 4858 | 3237 |
| FP | 3168 | 333 | 2462 | 863 | 342 |
| TN | 135316 | 138151 | 136022 | 137621 | 138142 |
| FN | 1730 | 1449 | 617 | 3026 | 4647 |
| Accuracy | 0.97 | 0.99 | 0.98 | 0.97 | 0.96 |
| Sensitivity | 0.78 | 0.82 | 0.92 | 0.62 | 0.41 |
| Specificity | 0.98 | 0.99 | 0.98 | 0.99 | 0.99 |
| PPV | 0.66 | 0.95 | 0.75 | 0.85 | 0.90 |
| NPV | 0.99 | 0.99 | 0.99 | 0.98 | 0.97 |
| F1 score | 0.72 | 0.88 | 0.83 | 0.71 | 0.56 |
| Cohen's Kappa | 0.70 | 0.87 | 0.81 | 0.70 | 0.55 |

Most false positives arose from a rather broad definition of “location” consistent across all libraries, which included also nouns related to physical locations such as “Stadt” (town, city), “Ort” (locality) or “Haus” (house) (Fig. S2). Only 26% of the location tokens were correctly identified as such by all libraries and 2% of the locations were missed by all libraries. Locations that were missed by all included Germanized spelling (e.g. “Hoschiarpur” for “Hoshiarpur”), latin spelling (e.g. “Centumcellae” for “Civitavecchia”), composite entities (e.g. “Gurjewscher Kreis”), historic regions (e.g. “Podolien”, “Gevaudan”) or ambiguous words (e.g. “Sind” is a location but also a conjugated verb form of “to be”). Compared to the other libraries, spaCy had a remarkable low number of FNs (Fig. S3). All libraries performed better on toponyms for cities or towns, whereas natural features or small villages proved to be more difficult (Fig. S4). spaCy identified historical regions correctly as toponyms more often than the other libraries (percentage false negatives among all historical regions: Spacy 4.8%, coreNLP 26.7%, Google 29.9%, Germaner 51.9% and geoparser 73.8%).

### Geocoding performance evaluation

To evaluate the geocoding performances, we used the 1856 unique location names from the plague dataset. Google and Geonames geolocated substantially more toponyms (74.8.% and 75.3%) than Geoparser.io (44.4%). Google and Geonames also geolocated more places correctly than geoparser (60.5% and 52.7% vs. 35.7%). Many of the mismatches occurred for regions where places were renamed as the ruling power changed, through colonization or the contraction and expansion of empires (e.g. in the regions of Armenia, Georgia or the Balkans) or where a phonetic translation of the original place name was used (e.g. entities located in Iran, Iraq, Ukraine, Russia or Kazakhstan) All geocoding services struggled to geocode historical regions, but Geonames and Google performed better (19.7% and 14.8% geocoded) than Geoparser (6.6%) (Fig. S4). Colloquial areas were also poorly geocoded (Google 23.5%, Geonames 20.6% and Geoparser 5.9%).

### Description of plague data sets

#### Comparison of Sticker and Biraben

The final Sticker dataset contained 4474 plague location observations, of which 91.4% could be localized exactly, 8.5% were localized approximatively and 0.1% could not be localized. Of the identified locations, 1631 were unique locations. The Biraben data set had much more data points and unique locations (11,180 observations, 2158 unique locations), of which 95.2% were localized exactly, 3.5% were localized approximately and 1.3% could not be localized) (Table S5). There was some overlap of the data points: 37% of the Sticker data were also in Biraben, and 15% of the Biraben data were also in Sticker. The majority of the data points in Sticker were located in Germany (13.6%), while Biraben had most data points in France (30.2%) (Fig. S5A). In both datasets, the majority of locations were places (Sticker 70.1%, Biraben 83.3%). Sticker contained more historical or colloquial regions or administrative units than Biraben, thus the average bounding box diagonal of a location was marginally larger for Sticker (17.6 km vs. 12.1 km) (Fig. S6). The most frequent places in Sticker were Istanbul (90 mentions, 2%), London (74 mentions, 1.7%) and Cairo (44 mentions, 1%) (Fig. S5B). Biraben listed the most outbreaks for London (166 mentions, 1.5%), Istanbul (118 mentions, 1.1%) and Algiers (114 mentions, 1%). Both data sets have the same overall temporal coverage from the Black Death period to the beginning of the 20^th^ century (Fig. S5C). However, the majority of entries in Biraben are from the 16^th^-17^th^ century, while the majority of Sticker is from the 17^th^-18^th^ century.

#### Spatial and temporal extent of second pandemic plague outbreaks

Fig. 2 shows all exactly localized places (without countries, regions or other administrative units) with plague outbreaks or occurrences reported during the second pandemic (1346-1894) resulting from both datasets. We found 1404 new observations (817 unique locations) in the Sticker dataset, which were not listed in Biraben. These were mainly in eastern Europe, southern Russia and the Caucasus region, as well as India and Iran. London had the largest number of outbreaks (265), followed by Istanbul/Constantinople (205), Algiers (138), Paris (115), Cairo (113), Izmir/Smyrna (107), Venice (103) and Amiens (102). As shown in Fig. 3, the spatio-temporal extent of plague outbreaks shifted considerably over time. Until the 17^th^ century we observe the majority of the data in Central Europe. In the 18^th^ century the focus appears to have shifted to Eastern Europe and North Africa. Finally, in the 19^th^ century the majority of outbreaks seemed to be reported in southeast Europe and West Asia.

**Fig. 2.**
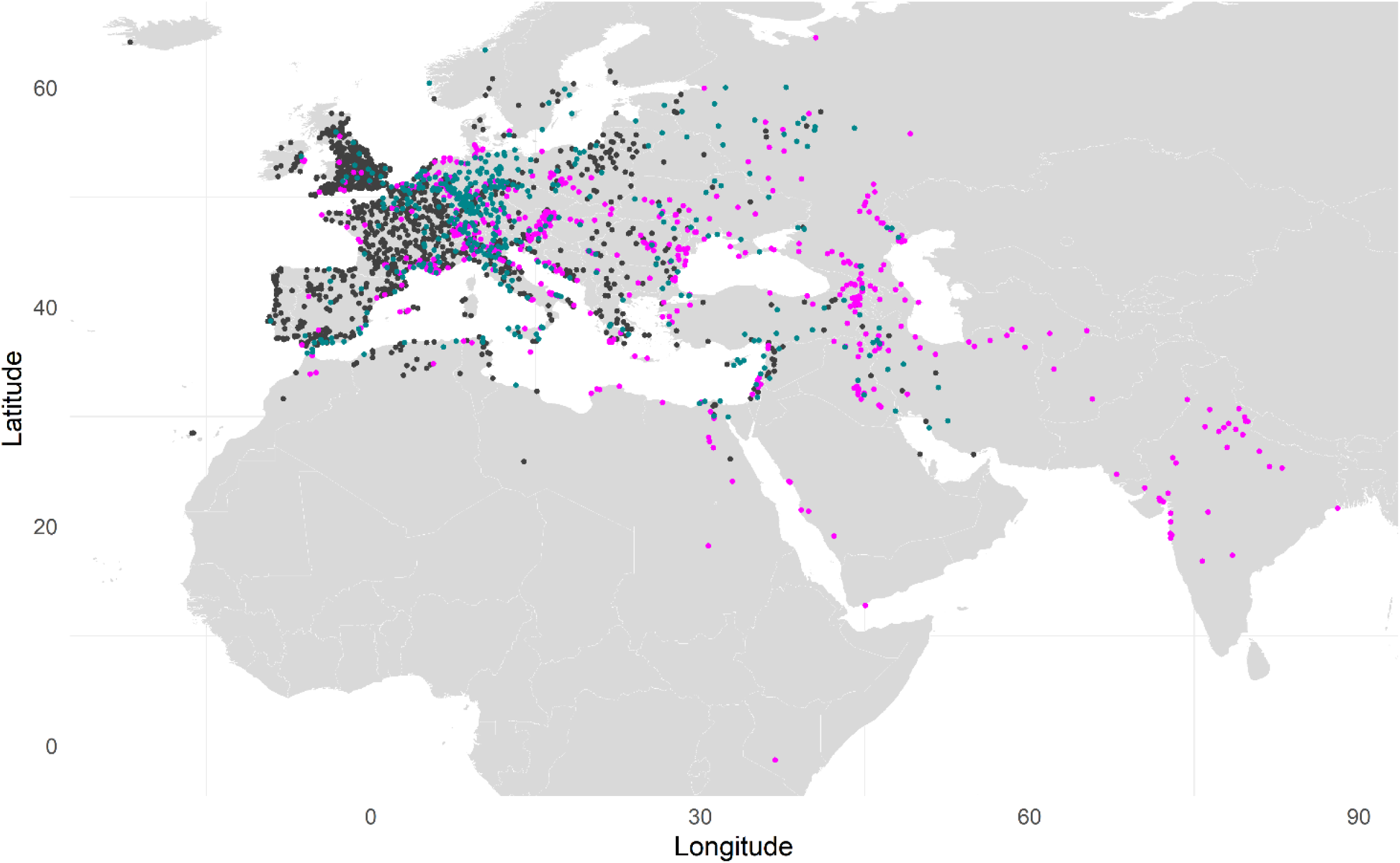
Places with plague during the second pandemic outbreaks in the data set of Biraben (grey), Sticker (pink) and both (blue).

**Fig. 3.**
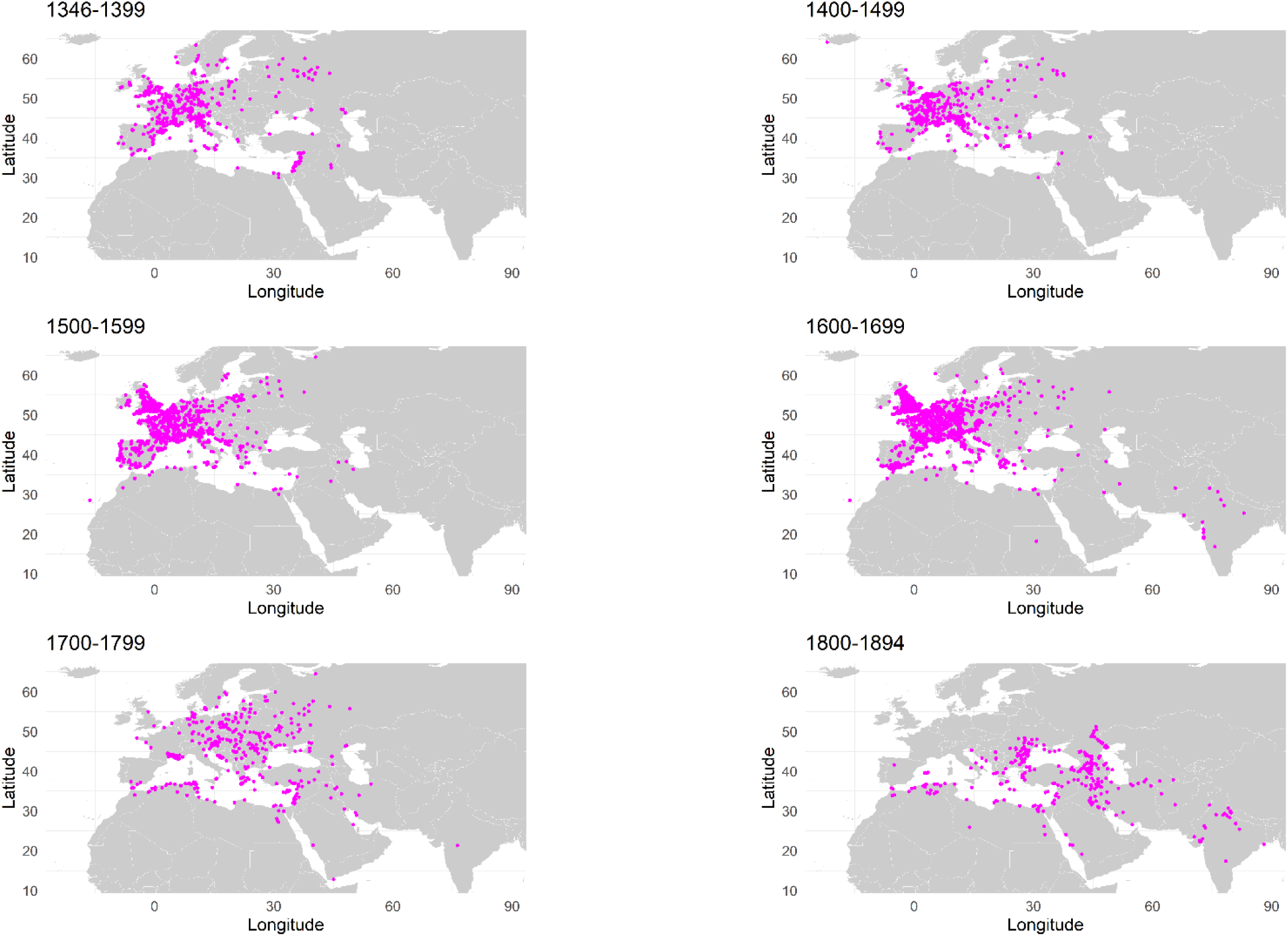
Spatio-temporal extent of the second pandemic plague outbreaks derived from Biraben and Sticker. The dots denote all exactly localized places, but exclude countries, regions or other administrative areas. For better readability, one data point in Nairobi (Kenya) of an outbreak in 1892 was omitted from the map.

## Discussion

We have demonstrated how natural language processing (NLP) libraries and geocoding/geoparsing tools can be used to detect, extract and georeference locations in a running text to facilitate the collection and digitization of plague data from a running text. We have shown that the performance of the different algorithms can vary substantially. For the given German text, Stanford’s coreNLP and spaCy had a better overall performance than Google’s NLP, germaNER and Geoparser.io. While spaCy was better at detecting the true locations (i.e. high sensitivity), Stanford coreNLP was marginally better at avoiding the non-locations (i.e. high specificity). However, all algorithms had a high specificity. Geoparser.io showed a poor performance and missed more than half of the true locations. According to the authors the algorithm works best with English texts, but there is limited information online on how the model was trained. It also showed a poorer performance in returning the correct coordinates compared to Google and Geonames. Overall, the sensitivity of all algorithms was imperfect, and a small proportion of locations remained undetected even with the best performing algorithm. All tested algorithms were substantially faster than manual annotation (less than 30 minutes vs. several days per annotator). The sensitivity of Stanford CoreNLP (0.81) and Google NLP (0.78) on Sticker’s treatise on plague was comparable to previous results from modern text corpora (0.64-0.89 and 0.77-0.87, respectively) (Dale, 2018; Gritta et al., 2020; Pinto et al., 2016; Schmitt et al., 2019), but spaCy outperformed its expectations, with a higher sensitivity (0.92) than advertised by the authors of the library (0.85) (Explosion, 2019a) and estimated in previous studies on English texts (0.57-0.75) (Gritta et al., 2020; Schmitt et al., 2019). Our F1 score for germaNER was somewhat lower (0.71) than evaluated by the authors of the algorithm (0.81) (Benikova et al., 2015). In terms of performance, it is more important to have a high sensitivity than a high specificity, because it is easier to remove false positives in the results than look for false negatives (missed locations) in a text. Thus, based on our findings, we recommend to use spaCy for entity recognition in combination with a geocoding services that also cover historical place names regions for the extraction of outbreak data from rather historical texts. All tested geocoding services showed a poor performance in geolocating historical regions and colloquial areas, but their performance could potentially be improved by passing on additional information such as the country or region to the geocoding service. Geonames also stores historical place names, and filtering all returned matches instead of accepting the best match could further improve the toponym recognition, but we have not tested this here.

### Advantages and limitations of NLP for the extraction of outbreak data

NLP libraries combined with geoparsers/geocoding tools are extremely useful to quickly generate quantitative data, but we have encountered some limitations for this specific project. As anticipated, these pre-trained models could not distinguish whether the mention of a geographical unit was related to a specific plague outbreak or not. This information can only be extracted from the context, but these standard models were not trained to recognize these situations. In this study, we have checked the link to a plague outbreak for each location entry manually, which is far from ideal. Moreover, the detection of time units was not optimal. We did not test the year numbers recognition formally, but we observed that Google, spaCy and Stanford CoreNLP don’t differentiate between years and any other number. For our plague dataset, we used regular expressions (regex), which can identify specific combinations of letters or numbers. The final linking of a specific year with a specific plague location was done manually again, since the order of appearance and the format in which years and locations were reported was not consistent throughout the text. Thus, the tested pre-trained NLP algorithms could not replace manual work entirely in our project.

The main potential of NLP and geoparsing for outbreak data extraction lies in custom trained models and reproducible, fully automated workflows. Some of the analyses that we did manually or in separate steps can potentially be improved with an automated procedure. Preprocessing of the raw OCR text prior to applying the NLP algorithms is inevitable, but OCR errors are often consistent and can be corrected with rule-based replacements as we have partially done here. NLP or geoparser libraries can be trained specifically on historical texts to improve the recognition of outdated spellings or old place names, and detect and extract relations of entities. The latter could potentially be used to link a specific outbreak to a specific place and time mention in the text. Text mining tools such as word embedding (i.e. linkage of entities by their proximity in the text) could also be used to detect relations. A custom trained model could also reduce the false positive rate for physical locations (e.g. “house” or “city”), which was an issue with all tested libraries. Examples of NLP models trained on epidemiological data include a recently published NLP pipeline (EpiTator) that uses the spaCy library in combination with Geonames specifically for the annotation of epidemiological data such as dates and date ranges, disease-related information and location data from running text (EcoHealth Alliance, 2019). This tool has also been custom trained on the incidence database of the Robert Koch Institut to detect emerging infections, and has shown a promising performance for country recognition (85% correctly classified), disease recognition (88% correctly classified) and date recognition (81% correctly identified) (Abbood et al., 2020). The aforementioned plague dot txt project is also pioneering the field with automated OCR optimization and extended NER for the recognition of plague-specific ontology and dates (Casey et al., 2020). Given the continuous emergence of infectious diseases and the exponentially increasing amount of epidemiological literature, we expect the landscape of NLP tools and pipelines trained on epidemiological texts to growth and improve in the coming years. Our dataset could be used by others as a training set for both improved toponym and relation recognition.

### Usage and limitations of geo-referenced plague datasets

We here also present two open, georeferenced plague datasets (Krauer and Schmid, 2021): the newly digitized Sticker dataset and an improved digitization of Biraben’s plague second pandemic appendix. The Biraben dataset has been digitized twice before (Atanasiu V et al., 2008; Buntgen et al., 2012), of which Büntgens version has been used by a number of studies (Schmid et al., 2015; Yue et al., 2016; Yue and Lee, 2018, 2020; Yue et al., 2017). These studies have rightfully drawn criticism for not contextualizing the biases and uncertainties inherent to such aggregated accounts that cover a vast amount of space and time (Roosen and Curtis, 2018; van Bavel et al., 2019). Both Biraben (and colleagues) as well as Sticker may have been more likely to include sources from specific regions or countries due to easier access to archives or familiarity with the language of the source texts. It is not by accident that the majority of plague occurrences of Biraben are in France and the majority of Sticker in Germany. Also, some regions might be poorly represented by sources due to cultural differences in what was perceived important to write down, or poor archiving conditions. These issues can lead to spatial and/or temporal selection bias in the data. Thus, the absence of plague occurrences listed in these datasets is not necessarily an absence of outbreaks. Moreover, the retrospective identification of a plague outbreak from historical sources is also often problematic, and the criteria that Sticker and Biraben used to include or exclude information are unclear. In this study, we have not verified the data, but we have provided references to the original sources wherever they were indicated by Sticker, which allows users to cross-check questionable entries. Biraben’s treatise was digitized from the tables provided in the appendix, which did not include references for each outbreak. However, the treatise itself includes an extensive bibliography for the origin of the data, which may be linked manually to specific outbreaks. Both datasets have inherent limitations due to the nature of the data collection and the digitization process. They are presented here as uncommented digitizations of all second and third plague pandemic entries provided in the Biraben and Sticker plague treatises, and should not be regarded as fully prepared and finalized plague data sets. Additional data cleaning and source verification is required depending on the research question and type of analysis. For example, the geographical scales of the observations in both datasets are very heterogeneous ranging from small villages to whole countries or historical regions spanning several hundreds of kilometers. For quantitative modelling studies, we recommend to work with data points that represent approximately the same geographic level. We have provided the bounding box coordinates and diagonal for each data point (which gives a rough estimate of the current geographical extent) as well as the type of location, which can be used to select data carefully. We also advise to check for duplicate entries for the same place in the same year, which occurred occasionally when the original dataset listed two separate entries for the same location (for example Saoudje and Boulak for Saoudje-Boulak, presently Mahabad, or individual parishes in London). As a caveat, most location coordinates provided in our datasets refer to the modern locations and we did not correct for potential geographical displacements over time. In the case of historical regions, we used the bounding coordinates based on historical maps on Wikipedia. The borders of these regions are thus only approximate.

Quantitative analyses will benefit from improved, georeferenced datasets, for example for the reconstruction of regional transmission chains or potentially the identification of putative historical plague reservoirs (Carmichael, 2014). As others have mentioned (Benedictow, 2019; van Bavel et al., 2019), data collections such as our compilation of Biraben and Sticker can act as a foundation to which more data are added (and faulty data are labelled as such) in order to build an updated database of global plague outbreaks. The growing number of scanned and OCR encoded documents made available online (for example on the Internet Archive) provides a rich resource for historical epidemiology, which should be used with the right tools and the necessary caution. Combining plague data from different sources to fill the spatial and temporal gaps could potentially reduce the problem of spatial and/or temporal representativeness, and improve our understanding of the spatio-temporal spread. Particularly, new data on the plague dissemination in neglected regions such as sub-Saharan Africa (Green, 2018), Turkey and Southern Asia (Green, 2014; Green, 2018; Varlik, 2020) could confirm whether the shift of plague activity from Europe to North Africa in the 16^th^ to 19^th^ century, and the growing presence of plague in Asia in the 17^th^ to 19^th^ century is a real pattern or merely an artefact of missing data in the centuries before. However, consistency in the data definition and collection is crucial. The understanding of the spatio-temporal dynamics of the past and present plague pandemic is a big challenge, which is best tackled with a collaborative and interdisciplinary effort, and in the spirit of open data.

## Data Availability

The R code and the digitised plague datasets are available in a public repository.

https://doi.org/10.5281/zenodo.6587267

## Author contributions

**Fabienne Krauer**: Conceptualization, Methodology, Software, Formal analysis, Data curation, Writing – Original draft, Writing – Review & Editing, Visualization **Boris V. Schmid**: Data curation, Writing – Review & Editing, Supervision, Funding acquisition

## Ethics statement

Not applicable.

## Data accessibility statement

The R code and the digitized plague datasets are available in a public repository (Krauer and Schmid, 2021) (https://doi.org/10.5281/zenodo.6587267).

