## Supplemental material for "Mapping the plague through natural language processing"

**Table S1.** Overview of works that compile places and times of historical plague outbreaks. This list of references is not exhaustive. Some of these works reference each other.

| Author | Publication | Geographical coverage | Temporal coverage | Reference |
|---|---|---|---|---|
| Villalba | 1803 | Spain | 1600-1801 | [1] |
| Hecker | 1865 | Europe | 14th century | [2] |
| Castaldi | 1872 | Kurdistan | 1871 | [3] |
| Tholozan | 1874 | Persia | 997-1871 | [4] |
| Martin | 1879 | Europe, Mediterranean | 1346-1700 | [5] |
| Radcliff | 1879 | Levante | 19th century | [6] |
| Creighton | 1891 | British Isles | 1348-1665 | [7] |
| Dörbeck | 1906 | Russia | 1352-1900 | [8] |
| Ferran | 1907 | Barcelona, ES | 1457-1652 | [9] |
| Gasquet | 1908 | Asia, Europe | 1348-1349 | [10] |
| Sticker | 1908 | Global | 1346-1819 | [11] |
| Sudhoff | 1923 | Germany, Bohemia, Austria | 15th century | [12] |
| Keyser | 1954 | Germany, Baltics | 1634-1640 | [13] |
| Bennassar | 1969 | North Spain | 1596-1599 | [14] |
| Revel | 1970 | Beneluxe | 1666-1670 | [15] |
| Shrewsbury | 1970 | UK | 1348-1665 | [16] |
| Biraben | 1975 | Europe, Mediterranean | 1346-1900 | [17] |
| Dols | 1977 | Middle East | 1349-1517 | [18] |
| Eckert | 1978 | Switzerland | 1563-1669 | [19] |
| Panzac | 1985 | Ottoman Empire | 1700-1850 | [20] |
| Alexander | 1986 | Russia | 1500-1800 | [21] |
| Noordegraaf | 1996 | The Netherlands | 1450-1668 | [22] |
| Congourdeau | 1999 | Constantinople | 1343-1466 | [23] |
| Panzac | 2004 | Greek Islands | 1686-1840 | [24] |
| Christakos | 2005 | Europe | 1347-1351 | [25] |
| Melikishvili | 2006 | Russia | 1401-1592 | [26] |
| Marien | 2009 | Ottoman Empire | 1347-1550 | [27] |
| Frandsen | 2010 | Baltic region | 1709-1730 | [28] |
| Varlik | 2015 | Ottoman Empire | 1350-1500 | [29] |
| Pribyl | 2017 | Eastern Anglia | 1348-1500 | [30] |
| Roosen | 2018 | Low Countries | 1349-1499 | [31] |
| Fazlinejad | 2018 | Iran | 14th and 15th century | [32] |

### Supplemental methods

#### Text S1.

#### Source text

Our source text is a German plague treatise ("Geschichte der Pest") published in 1908 by Georg Sticker [11]. Sticker was a German physician, who together with Robert Koch was sent to Bombay to investigate the plague epidemic in 1897. In the first part of the book, he narrates the historical spread of plague on five continents (Europe, Asia, the Americas, Africa and Australia) in chronological order from biblical times to 1908. In the second part, Sticker elaborates on plague ecology, microbiology and pathology. For the historical part, he combined information from secondary literature, but he also consulted original sources (see his introduction). It is divided in 16 Chapters, of which the first four chapters are about the first pandemic, chapters 5 to 15 are about the second pandemic and the chapters 15 and 16 refer to the third pandemic. For our analysis, we focus on plague during the second and third pandemic, corresponding to the chapters five to sixteen of Sticker's treatise on plague (pages 42 to 399).

**Fig. S1.** Schematic workflow of our analysis.

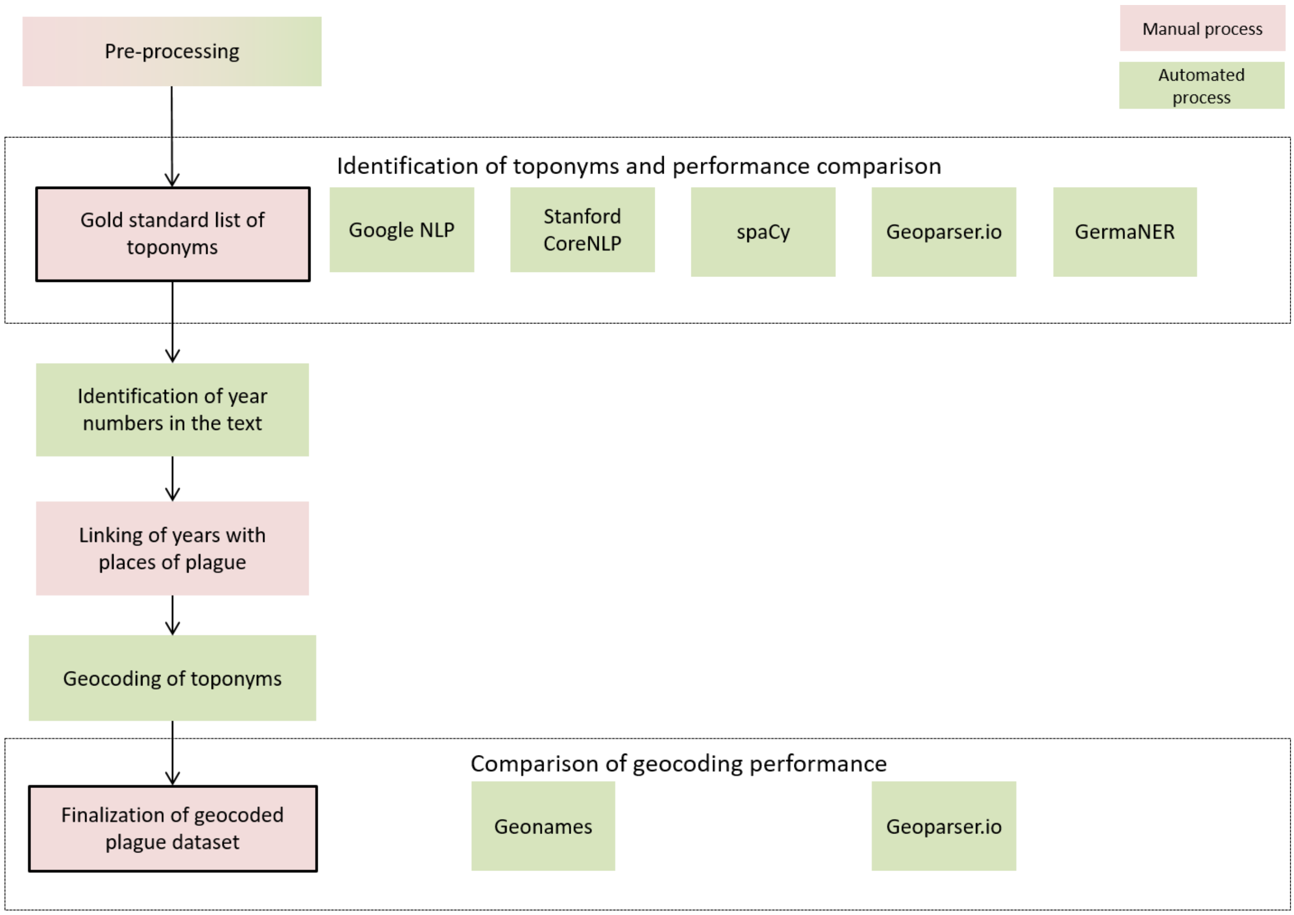

**Table S2.** Technical comparison of the five NLP algorithms tested. All studied tools are based on machine learning algorithms. CNN=convoluted neural network, CRF=conditional random field.

| | **spaCy** | **Google NLP** | **Stanford CoreNLP** | **GermaNER** | **Geoparser** |
|---|---|---|---|---|---|
| **model type** | CNN | CNN | CRF | CRF | ? |
| **Custom model** | yes | yes (Google AutoML) | yes | yes | no |
| **open source** | yes | no | yes | yes | no |
| **pricing** | free | free up to 5,000,000 characters/month | free | free | free up to 1000 API calls/month |
| **pro-graming language** | Python/Cython | ? | Java | Java | ? |
| **R package** | spacyr | | coreNLP | | |
| **supported languages** | en, cn, dk, nl, en, fr, de | en, cn, dk, nl, en, fr, de, es, jp, it, korean, pt, ru | en, de, es, cn | de | en |
| **training data set** | TIGER, WikiNER | ? | CoNLL 2003 | GermEval 2014 NoSta-D Named Entity | ? |
| **tasks** | tokenization, POS, NER | tokenization, POS, NER | tokenization, POS, NER | NER | NER LOC and geocoding |
| **issues** | | POS and NER require two steps, tokenization is not identical. Umlaut must be queried as UTF-16. | Umlaut returned incorrectly with coreNLP | | |
| **author(s)** | Explosion AI | Google Inc. | Finkel et al | Benikova et al | Geoparser Inc. |
| **license** | MIT | | GNU v3 | ASL 2.0 | |

**Text S2.**

**Definition of performance indicators**

The accuracy is the overall proportion of correct predictions (positive and negative) among all predictions. The sensitivity (recall) is a measure of how good the algorithm is at correctly detecting locations. The specificity (selectivity) estimates the probability of correctly detecting a non-location. The positive predictive value (PPV or precision) is the proportion of predicted locations that are true locations. The negative predictive value (NPV) does the inverse, i.e. it estimates the probability that if a token is identified as a non-location, it is a true non-location. Of note, PPV and NPV depend on the proportion of locations in the text, i.e. they are intrinsic and cannot be compared between different texts. The F1 score is a harmonic average of sensitivity and precision with 0 being the lowest performance and 1 being the perfect performance. Finally, Cohen's Kappa coefficient measures the observed accuracy compared to the expected accuracy when all agreement is by chance (random). A coefficient of 1 indicates perfect agreement; a coefficient of 0 indicates no agreement.

**Table S3.** Definitions of the performance measures used in the main analysis. TP=true positives, TN=true negatives, FP=false positives, FN=false negatives.

| Measure | Definition |
| --- | --- |
| Accuracy | $\frac{\mathrm{TP} + \mathrm{TN}}{\mathrm{TP} + \mathrm{TN} + \mathrm{FP} + \mathrm{FN}}$ |
| Sensitivity (Recall, true positive rate) | $\frac{\mathrm{TP}}{\mathrm{TP} + \mathrm{FN}}$ |
| Specificity (Selectivity, true negative rate) | $\frac{\mathrm{TN}}{\mathrm{TN} + \mathrm{FP}}$ |
| Positive predictive value (PPV) | $\frac{\mathrm{TP}}{\mathrm{TP} + \mathrm{FP}}$ |
| Negative predictive value (NPV) | $\frac{\mathrm{TN}}{\mathrm{TN} + \mathrm{FN}}$ |
| F1 score | $2 * \frac{\mathrm{PPV} * \mathrm{sensitivity}}{\mathrm{PPV} + \mathrm{sensitivity}}$ |
| Cohen's Kappa | $\frac{\mathrm{accuracy} - \mathrm{p_e}}{1 - \mathrm{p_e}}$<br>where<br>pe (probability of agreement by chance) =<br>$\frac{\frac{(\mathrm{TP} + \mathrm{FN}) * (\mathrm{TP} + \mathrm{FP})}{\mathrm{TP} + \mathrm{TN} + \mathrm{FP} + \mathrm{FN}} + \frac{(\mathrm{TN} + \mathrm{FN}) * (\mathrm{FN} + \mathrm{FP})}{\mathrm{TP} + \mathrm{TN} + \mathrm{FP} + \mathrm{FN}}}{\mathrm{TP} + \mathrm{TN} + \mathrm{FP} + \mathrm{FN}}$ |

**Text S3.**

**Re-digitization of Biraben's plague list**

In 1975, the French Jean-Noël Biraben published one of the most popular treatises about plague outbreaks in Europe and the Mediterranean during the first and second pandemic [17]. With the help of dozens of other researchers, he consulted more than one thousand studies and reports and synthesized the information in two books including a large time series dataset with years and place names in the appendix [17]. Earlier historians such as Hecker [2], Martin [5] and Sticker [11] laid the groundwork with their narrative accounts of places and countries that had plague outbreaks in specific years, but it was Biraben who compiled a large amount of information in a tabular format. In the appendix of his work, he provided two lists of places or regions that reported plague in a specific year. The first table consisted of three pages addressing plague outbreaks during the First plague pandemic (between 541 – 775), followed by 73 pages addressing plague outbreaks during the Second Plague Pandemic. Both tables are grouped into different chapters according to their geographical region. The book also includes a bibliography, which the work is based on, but individual place names are not linked to a specific source. His dataset has been widely used and criticized by historians and scientists. For example Roosen and Curtis [33] mention the problem of overrepresentation of specific countries such as France or England or the underrepresentation of some areas such as the Low countries, respectively. Despite its methodological flaws, it is the best available dataset as of today for quantitative studies about the dissemination of plague in Europe during the second pandemic.

There have been two attempts to digitize and geocode the Biraben collection. The first dataset was generated by the EU-funded 'project Bernstein', which digitized historic documents and made them available in an electronic database [34]. The dataset contains 5837 observations (949 unique locations) with information about the year, the status ("region" or "location"), the modern place name, lower level administrative area names, the country name as well as the coordinates (lat/lon in WGS84). However, the original place names have not been preserved completely, which makes it difficult to compare it to other datasets or link it with the raw data. This dataset is publicly available on the project website, but it was not formally published in a peer-reviewed journal. The second dataset was digitized by Büntgen et al and was published in 2012 as a correspondence in Clinical Infectious Diseases [35]. This dataset contains 6929 observations (1171 unique locations) with information on the modern place name, the year and the coordinates. The original place names are missing as well and there is no country information. Both digitized versions are a subset of the original data set: The Büntgen data set lacks the entries for Scandinavia, the Balkans, the Beneluxe and parts of the Levante, the Bernstein dataset lacks the entries for the Maghreb, the Levante, the Southern Balkans and generally all data after the year 1600. We therefore sought to re-digitize and geocode the complete appendix four of Biraben and to provide an updated, improved digital version of Biraben's data set.

We scanned the complete appendix 4, performed optical character recognition (OCR) using Adobe Acrobat Pro and produces a plain text file. We manually corrected misaligned text and unreadable characters. The file was then read into R and a raw dataset with all observations was created. We preserved Biraben's distinction between a region and a place ("type_orig"), whether there were doubts about a place having plague ("certain") and in which chapter the entry occurs ("chapter"). We then batch-geocoded all locations using the methodology described in the main text. We searched only countries covered by the corresponding chapter given by Biraben, which increased the accuracy of the matches. Unclear or ambiguous locations were checked by consulting some of the original literature used by Biraben. As for the Sticker dataset, places which could not be localized exactly (including instances were Biraben described as place as “environs X”) were geocoded according to the next lower identifiable level administrative unit and marked as “approximate”. Places that could not be localized at all were marked as “unknown”. For London, Biraben occasionally lists both the city name as well as individual parishes affected by plague for a given year. This leads potentially to an inflation of data and was addressed by geocoding any parish belonging to the city of London within and without the wall (situation as of 1870) as “London”. This resulted in multiple entries for London for the same year. Researchers should apply some data cleaning or filtering before using our dataset.

### Supplemental results

**Table S4.** Comparison of identified results of tokens and entities by five different NLP and geoparser libraries.

| | Gold standard | Google NLP | Stanford CoreNLP | spaCy | germaNER | Geoparser |
|---|---|---|---|---|---|---|
| Benchmark | ~ 1 week | <1 min (POS)<br><1 min (NER) | 28 mins | <1 min | 11 mins | <1 min |
| Tokens recognized | 146,368 | 146,340 | 146,743 | 146,767 | * | * |
| Entities classified N (%) | | | | | | |
| Total | 7884 | 33,925 (100) | 9522 (100) | 12,963 (100) | 50,374 | 3563 |
| Location | 7884 | 9246 (27.25) | 6989 (73.40) | 10,050 (77.53) | 5885 (11.68) | 3563 |
| Consumer good | | 765 (2.25) | | | | |
| Event | | 2325 (6.85) | | | | |
| Organization | | 850 (2.51) | 240 (2.55) | 250 (1.93) | 246 (0.49) | |
| Other / Misc | | 14,875 (43.85) | 817 (8.52) | 1047 (8.08) | 43,843 (87.03) | |
| Person | | 5755 (16.96) | 1476 (15.53) | 1616 (12.47) | 400 (0.79) | |
| Work of Art | | 109 (0.32) | | | | |
| N tokens after mapping (%) | | | | | | |
| Location | | 9322 (6.4) | 6768 (4.6) | 9729 (6.6) | 5721 (3.9) | 3579 (2.4) |
| Other / Not recognized | | 137046 (93.6) | 139600 (95.4) | 136639 (93.4) | 140647 (96.1) | 142789 (97.6) |

*GermaNER and Geoparser.io don’t return a tokenization. Geoparser.io returns only tokens that were recognized as a toponym. GermaNER requires an a-priori tokenization of the text, we here used the tokenization of spaCy, which was the most stringent.

**Fig. S2.** Frequency of false positives by NLP library. For clarity, we include only tokens which appear more than 10 times throughout the text.

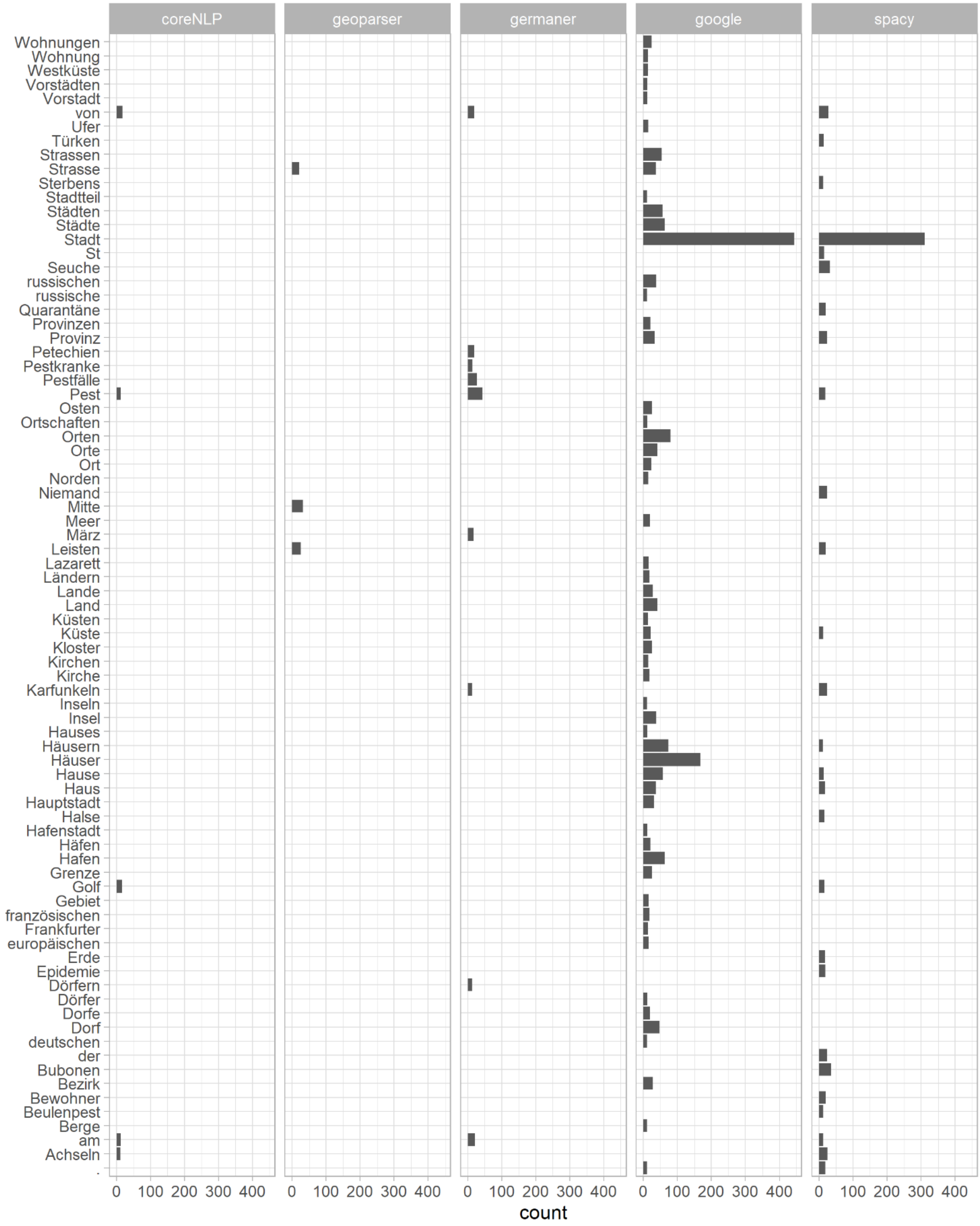

**Fig. S3.** Frequency of false negatives by NLP library. For clarity, we include only tokens which appear more than 5 times throughout the text. Geoparser was omitted from the graph due to its very high number of false negatives, which rendered the figure unreadable.

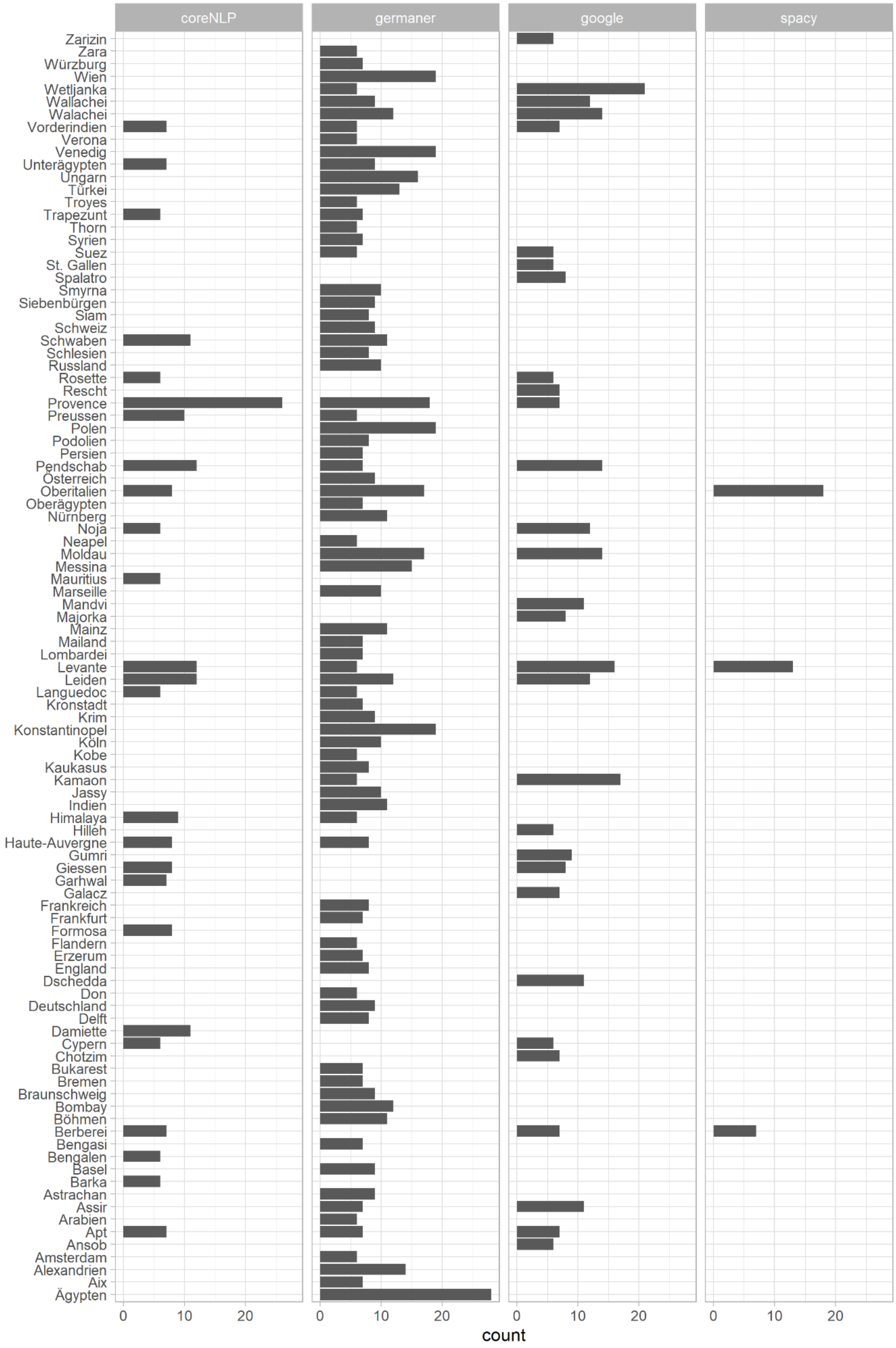

**Fig. S4.** Percentage of false negative (FN) toponyms by NLP library and type. The number of toponyms in total is given in parenthesis for each type next to the label. The denominator corresponds to the total number of toponyms, the numerator corresponds to the number of false negatives. The graph includes only tokens which were listed as plague locations (the type of geographical entity is not available for the full gold standard list of toponyms, only for the geocoded plague dataset).

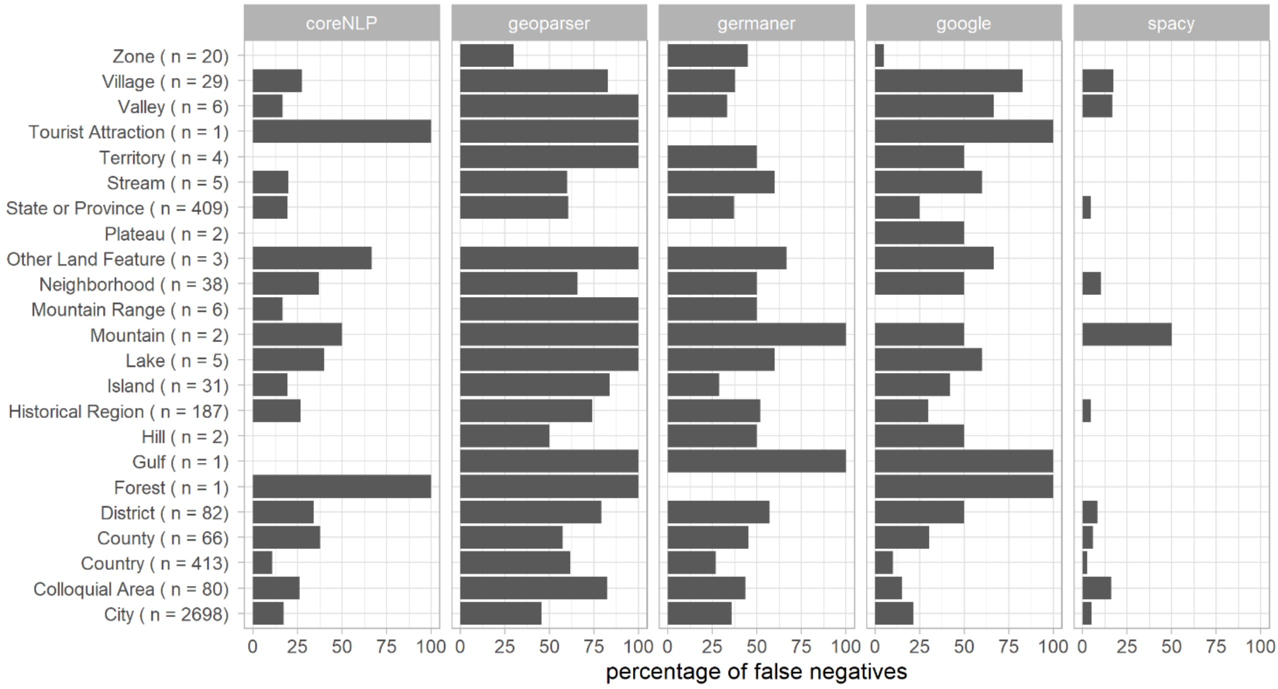

**Fig. S5.** Percentage of correctly geocoded toponyms by NLP library and type. The denominator corresponds to the total number of toponyms for each type, the numerator corresponds to the number of correctly geocoded toponyms. The graph includes only tokens which were listed as plague locations (the type of geographical entity is not available for the full gold standard list of toponyms, only for the geocoded plague dataset).

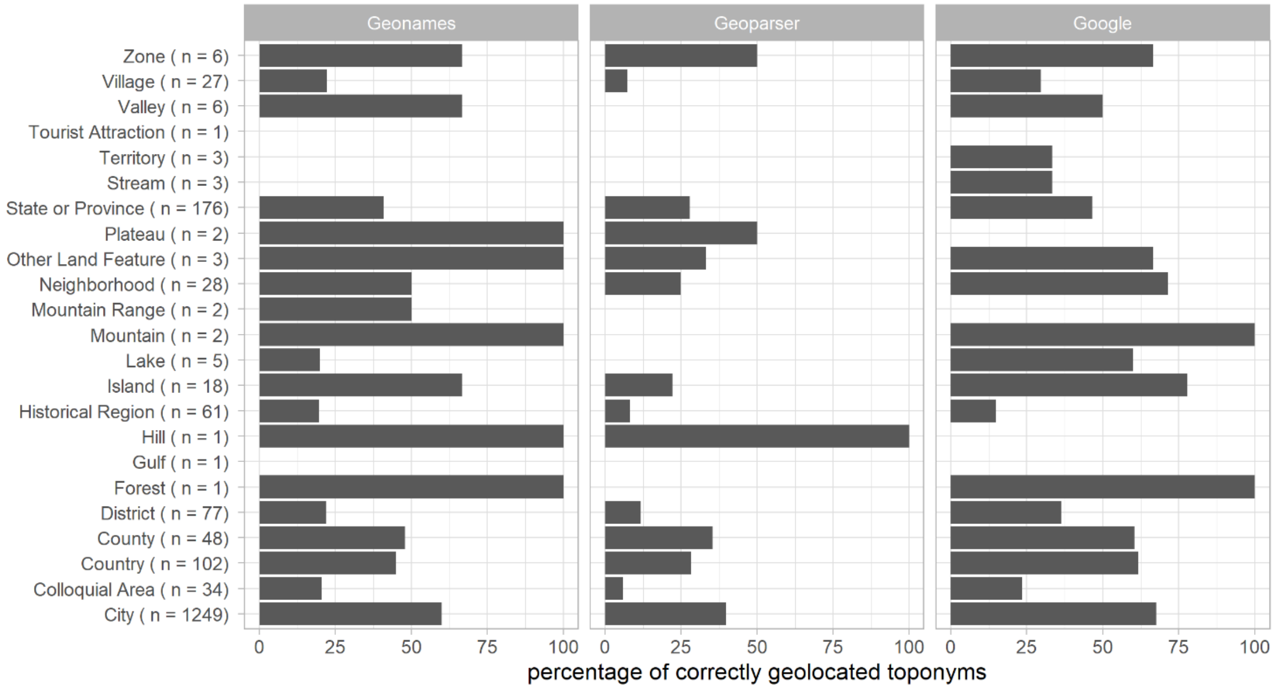

**Table S5.** Comparison between the Sticker and the Biraben dataset.

| | Sticker | Biraben |
|---|---|---|
| N total locations | 4474 | 11180 |
| N exactly identified locations | 4087 | 10644 |
| N approximate locations | 379 | 392 |
| N unidentified location | 8 | 144 |
| N unique locations (exact & approximate) | 1631 | 2158 |
| Spatial coverage | | |
| Latitude | -38°N to 75° N | 12° N to 68° N |
| Longitude | -158 ° E to 166 ° E | -22 °E to 97° E |
| N countries | 104 | 65 |
| Types (% of total) | | |
| City | 66.5 | 80 |
| Country | 10.1 | 4.6 |
| State or Province | 9.9 | 5.3 |
| Historical Region | 4.7 | 2.7 |
| District | 2 | 2.5 |
| Colloquial Area | 1.8 | 1.2 |
| County | 1.5 | 1.3 |
| Neighborhood | 0.9 | 0.2 |
| Island | 0.7 | 1 |
| Village | 0.7 | 0.4 |
| Zone | 0.5 | 0.4 |
| Lake | 0.1 | 0 |
| Mountain Range | 0.1 | 0.1 |
| Other Land Feature | 0.1 | 0 |
| Stream | 0.1 | 0.2 |
| Territory | 0.1 | 0 |
| Valley | 0.1 | 0 |
| Forest | 0 | 0 |
| Gulf | 0 | 0 |
| Hill | 0 | 0 |
| Mountain | 0 | 0.1 |
| Plateau | 0 | 0 |
| Tourist Attraction | 0 | 0 |
| Park | 0 | 0 |
| Ruin | 0 | 0 |
| Community | 0 | 0.1 |
| Municipality | 0 | 0.1 |
| Years (range) | 1346-1908 | 1346-1900 |
| Century (% of total) | | |
| $14^{th}$ | 10.9 | 9.9 |
| $15^{th}$ | 6.9 | 15.8 |
| $16^{th}$ | 12 | 31.1 |
| $17^{th}$ | 23.4 | 30.3 |
| $18^{th}$ | 17.8 | 8.2 |
| $19^{th}$ | 18.8 | 4.7 |
| $20^{th}$ | 10.3 | 0 |

**Figure S5.** Comparison of the spatial and temporal coverage of the Sticker and Biraben dataset. (A) Relative frequency of countries (as percentage of total observations), (B) Absolute frequency of most often mentioned places, (C) yearly number of locations reported to have had a plague outbreak.

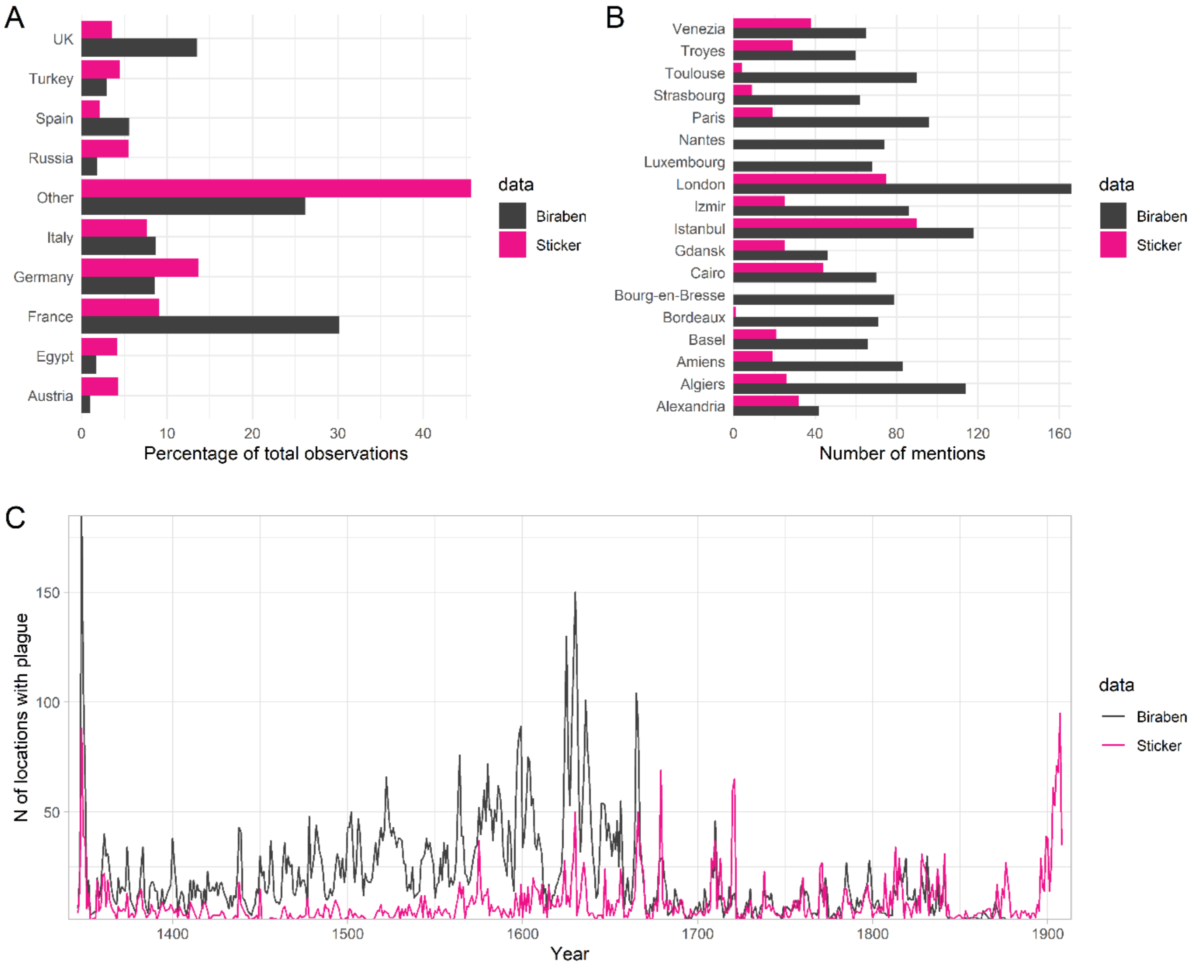

**Figure S6.** Diagonal of the bounding box in km according to type and dataset.

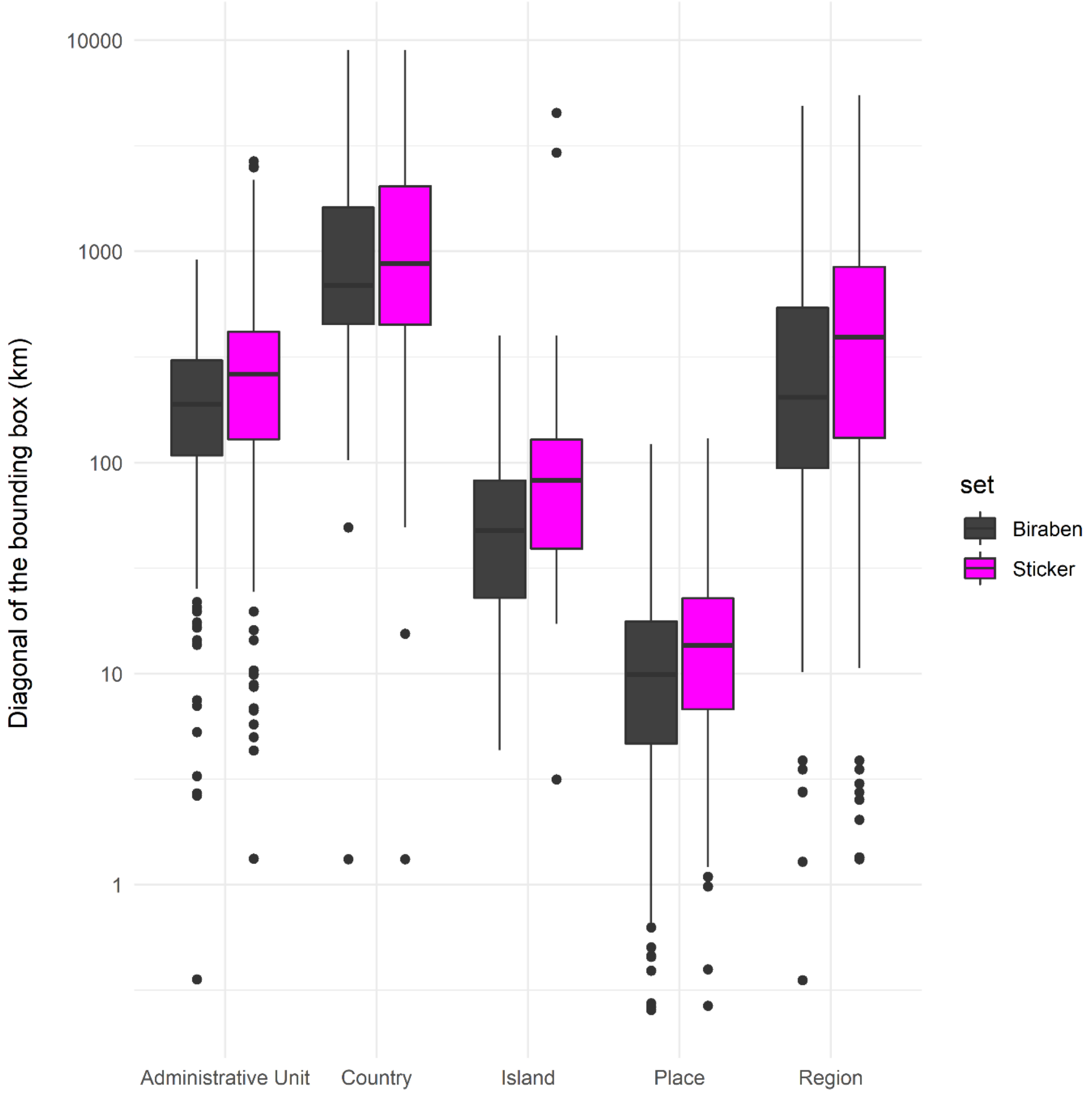

### References


[1] Villalba, J.d. 1802 *Epidemiologia española, o, Historia cronologica de la pestes, contagios, epidemias y epizootias : que han acaecido en España desde la venida de los cartagineses, hasta el año 1801 : con noticia de algunas otras enfermedades de esta especie que han sufrido los españoles en otros reynos, y de los autores nacionales que han escrito sobre esta materia, asi en la peninsula como fuera de ella*, Madrid : En la Imprenta de Don Mateo Repullés.

[2] Hecker, J.F.C. 1865 *Die grossen Volkskrankheiten des Mittelalters: Historisch-pathologische Untersuchungen. Gesammelt und in erweiterter Bearbeitung*. Berlin, Enslin.

[3] Castaldi, M. 1872 *La peste dans le Kurdistan persan. Rapport addressé au conseil supérieur de santé à Constantinople*.

[4] Tholozan, J.D. 1874 *Histoire de la peste bubonique en Perse, ou détermination de son origine, de sa marche, du cycle de ses apparitions et de la cause de sa prompte extinction*. Paris, G. Masson.

[5] Martin, C. 1879 Versuch einer geographischen Darstellung einiger Pestepidemien. *Dr. A. Petermann's Mittheilungen aus Justus Perthes' Geographischer Anstalt* **XXV**.

[6] Radcliffe, N. 1879 Papers relating to the modern history and recent progress of Levantine plague. . (London.

[7] Creighton, C. 1891 *A history of epidemics in Britain*. 2nd ed. Cambridge, Cambridge University Press.

[8] Dörbeck, F. 1906 *Geschichte der Pestepidemien in Russland von der Gründung des Reiches bis auf die Gegenwart*. Breslau, Kern.

[9] Ferrán i Clua, J., Viñas y Cusí, F. & de Grau, C.R. 1907 *La Peste Bubónica: memoria sobre la epidemia ocurrida en Porto en 1899*. Barcelona, Typografia Sucesor F. Sanchez.

[10] Gasquet, F.A. 1908 *The Black Death of 1348 and 1349*, G. Bell.

[11] Sticker, G. 1908 *Abhandlungen aus der Seuchengeschichte und Seuchenlehre. Band 1: Die Pest*. Giessen, A. Töpelmann.

[12] Sudhoff, K. 1922 Pestschriften aus den ersten 150 Jahren nach der Epidemie des „schwarzen Todes“ 1348. XV. Pesttraktate aus dem östlichen Süddeutschland, Böhmen und Österreich in der 2. Hälfte des 15. Jahrhunderts. *Archiv für Geschichte der Medizin* **14**, 1-25.

[13] Keyser, E. 1954 **Die Ausbreitung der Pest in den deutschen Städten**. In: *Ergebnisse und Probleme moderner geographischer Forschung. Festschrift für Hans Mortensen.* (Bremen, Dorn.

[14] Bennassar, B. 1969 *Recherches sur les grandes épidémies dans le nord de l'Espagne à la fin du XVIe siècle : problèmes de documentation et de méthode*. Paris, S.E.V.P.E.N.

[15] Revel, J. 1970 Autour d'une épidémie ancienne : la peste de 1666-1670. *Revue d’histoire moderne et contemporaine* **17**, 953-983. (doi:10.3406/rhmc.1970.2125).

[16] Shrewsbury, J.F.D. 1970 *A History of Bubonic Plague in the British Isles*. Cambridge, Cambridge University Press.

[17] Biraben, J.-N. 1975 *Les hommes et la peste en France et dans les pays européens et méditerranéens*. Paris, Mouton.

[18] Dols, M.W. 1977 *The "black death" in the Middle East*. Princeton (N.J.), Princeton University Press.

[19] Eckert, E.A. 1978 *Boundary formation and diffusion of plague : Swiss epidemics from 1562 to 1669*.

[20] Panzac, D. 1985 *La peste dans l'empire ottoman, 1700-1850*, Peeters.

[21] Alexander, J.T. 1986 Reconsiderations on plague in early modern Russia, 1500-1800. *Jahrb Gesch Osteur* **34**, 244-254.

[22] Noordegraaf, L. & Valk, G. 1996 *De gave Gods: de pest in Holland vanaf de late middeleeuwen*, B. Bakker.

[23] Congourdeau, M.H. 1999 [Black Death in Constantinople (1343-1466]. *Med Secoli* **11**, 377-389.

[24] Panzac, D. 2004 La peste dans les possessions insulaires du Grand Seigneur (xviie-xixe siècles). In: *Insularités ottomanes* (eds. N. Vatin & G. Veinstein), pp. 223-240, Institut français d'études anatoliennes.
[25] Christakos, G., Olea, R., Serre, M., Yu, H. & Wang, L. 2005 *Interdisciplinary Public Health Reasoning and Epidemic Modelling: The Case of Black Death*. Berlin, Heidelberg, Springer.
[26] Melikishvili, A. 2006 Genesis of the anti-plague system: the Tsarist period. *Crit Rev Microbiol* **32**, 19-31. (doi:10.1080/10408410500496763).
[27] Marien, G. 2009 The Black Death in early Ottoman territories: 1347–1550 [PhD dissertation], Bilkent University, Ankara, Turkey.
[28] Frandsen, K.E. 2010 *The Last Plague in the Baltic Region, 1709-1713*, Museum Tusculanum Press; 537 p.
[29] Varlik, N. 2015 *Plague and Empire in the Early Modern Mediterranean World*. Cambridge, Cambridge University Press.
[30] Pribyl, K. 2017 Climate and the Plague, 1348–1500. In: *Farming, Famine and Plague* (pp. 199-223.
[31] Roosen, J. & Curtis, D.R. 2018 The 'light touch' of the Black Death in the Southern Netherlands: an urban trick? *Econ Hist Rev* **72**, 32-56. (doi:10.1111/ehr.12667).
[32] Fazlinejad, A. & Ahmadi, F. 2018 The Black Death in Iran, according to Iranian Historical Accounts from the Fourteenth through Fifteenth Centuries. *Journal of Persianate Studies* **11**, 56-71. (doi:10.1163/18747167-12341321).
[33] Roosen, J. & Curtis, D.R. 2018 Dangers of Noncritical Use of Historical Plague Data. *Emerging infectious diseases* **24**, 103-110. (doi:10.3201/eid2401.170477).
[34] Atanasiu V, Priol C, Tournieroux A & E, O. Georeferences for places of plague occurrence in Europe 1347-1600. Available from: https://bernstein.oeaw.ac.at/atlas/yersinia-description.pdf. [Accessed Mar 3, 2019].
[35] Buntgen, U., Ginzler, C., Esper, J., Tegel, W. & McMichael, A.J. 2012 Digitizing historical plague. *Clin Infect Dis* **55**, 1586-1588. (doi:10.1093/cid/cis723).
